# Cohort Study: The Accuracy of Screening Methods of COVID-19 in Pregnancy: Practical Approach in Low Resources Health Services

**DOI:** 10.1101/2021.09.20.21263866

**Authors:** Muhammad Ilham Aldika Akbar, Khanisyah Erza Gumilar, Eccita Rahestyningtyas, Manggala Pasca Wardhana, Pungky Mulawardhana, Jimmy Yanuar Anas, Ernawati, Muhammad Ardian Cahya Laksana, Hermanto Tri Joewono

**Affiliations:** Department Obstetrics & Gynaecology, Faculty of Medicine Universitas Airlangga - Universitas Airlangga Academic Hospital, Kampus A UNAIR, Jl. Mayjen Prof. Dr. Moestopo No. 47, Surabaya, Indonesia; Department Obstetrics & Gynaecology, Faculty of Medicine Dr. Soetomo General Academic Hospital, Universitas Airlangga Hospital, Jl. Dharmahusada Permai, Mulyorejo, Kec. Mulyorejo, Kota SBY, Jawa Timur 60115, Surabaya, Indonesia

**Keywords:** COVID-19, Pregnancy, Screening, Diagnosis

## Abstract

**Background:** All pregnant women in labor should be universally screened for Coronavirus Disease 2019 (COVID-19) during pandemic periods using *Reverse Transcriptase Polymerase Chain Reaction* (RT-PCR) test. In many low-middle income countries, screening method was developed as an initial examination because of limited availability of RT-PCR tests.

**Objectives:** This study aims to evaluate the screening methods of COVID-19 accuracy in pregnant women.

**Material and Methods:** We recruited all pregnant women with suspicion of COVID-19 from April - August 2020 at Universitas Airlangga hospital, Surabaya, Indonesia. The participant was divided into two groups based on RT-PCR results: COVID-19 and non-COVID-19 group. The proportion of positive signs & symptoms, rapid antibody test, abnormal findings in chest x-ray, and neutrophil to lymphocyte ratio (NLR) value were then compared between both groups. The sensitivity, specificity, positive predictive value (PPV), negative predictive values (NPV), and diagnostic accuracy (DOR) were calculated.

**Results:** A total 141 pregnant women with suspected COVID-19 cases were recruited for this study. This consist of 62 COVID-19 cases (43.9%) and 79 non COVID-19 pregnant women (56.1%). The sensitivity, spesificity, PPV, NPV, and diagnostic accuracy of each parameter are as follow: clinical sign & symptoms (24.19%, 75.95%, 3.92%, 96.11%, 65.87%), rapid antibody test (72.73%, 35.06%, 4.35%, 96.94%, 36.53%), chest x-ray (40.68%, 59.45%, 3.92%, 96.11%, 58.76%), and NLR > 5.8 (41.38%, 72%, 5.66%, 96.80%, 70.81%).

**Conclusions:** The use of combined screening methods can classify pregnant women with high-risk COVID-19 before definitively diagnosed with RT-PCR. This practice will help to reduce RT-PCR need in a limited resources country.

## Introduction

From the start of March 2020 until now (January 12th, 2021), the Coronavirus Disease 2019 (COVID-19) pandemic has infected 91.314.370 peoples worldwide and lead to 1.952.879 total death (1). Indonesia has become one of the most important spreading centers in South Asia, especially Southeast Asia. Until now (January 12th, 2021), there has been a total of 828.000 COVID-19 cases in Indonesia, with total death of 24.129 peoples (Case Fatality Rate [CFR] 2.91%) (2). COVID-19 pandemic also has a significant impact on pregnancy, which is the increased maternal morbidity and mortality. Pregnancy with COVID-19 poses specific challenges because most of the cases were asymptomatic. A study performed at Universitas Airlangga Hospital found 75.8% of pregnancies with confirmed COVID-19 have no signs or symptoms. This finding is in line with the New York study, which found 87.9% of pregnant women with COVID-19 also asymptomatic (3). It is essential to determine the COVID-19 status of pregnant or delivering women because confirmed cases need unique and more comprehensive management. Many international guidelines from Obstetrics & Gynaecologic organization recommend universal screening of COVID-19 for women in labor during this pandemic period (4–9). In the developed countries with adequate resources, universal screening of COVID-19 is performed using a gold standard: *Reverse Transcriptase Polymerase Chain Reaction* (RT-PCR) from the oropharynx and nasopharyngeal swab samples.

However, in low-middle income countries without sufficient resources and limited RT-PCR availability for diagnostic, such practice is difficult to perform. Therefore some modification was made to overcome this problem. The golden standard of RT-PCR is only performed in a case with a high suspicion of COVID-19 (high-risk group). Initial classification of a patient is performed using combined screening methods, including sign & symptoms, physical examination, laboratory result, radiology examination (Chest CT-scan, Chest X-ray), and serological test (*rapid test antibody*). The clinical manifestation of COVID-19 includes fever, cough, dyspnea, myalgia, sore throat, anosmia, ageusia, dan nausea vomiting (10–12). These symptoms become the most basic screening methods for all women in labor. The laboratory examination usually shows a sign of lymphocytopenia, increased C-reactive protein (CRP), leukocytosis, lymphopenia, neutrophilia, and increased neutrophil/lymphocyte ratio (NLR) (13,14). Predominant findings of peripheral airspace shadowing and bilateral, multi-lobar ground-glass opacities or consolidation in the chest x-ray or chest CT indicating a suspicion of COVID-19 infection. (15). A serological test in low-middle income countries is used as a additional test to diagnose Coronavirus disease, besides the ideal molecular viral test. The advantages of rapid antibody tests include easy to perform, cheap, and does not need a complete facility or trained examiner compare to the RT-PCR examination (16).

Considering the limited availability of diagnostic RT-PCR of COVID-19 in many hospitals, we conduct a study to assess screening methods’ diagnostic accuracy in pregnancy. This study evaluates the diagnostic accuracy of screening methods of COVID-19 on pregnancy by comparing it with the RT-PCR as the gold standard.

## Material and Methods

This study is part of a cohort study of suspected COVID-19 cases in pregnant women in Universitas Airlangga Hospital (Surabaya, Indonesia) from April - August 2020. This study aims to evaluate the accuracy of our screening methods of COVID-19 in pregnant women. We performed a universal screening COVID-19 in pregnant women in labor based on our original screening methods: which is the presence of one of the clinical sign-symptoms of covid 19 (fever, cough, dyspnea, sore throat, myalgia, or nausea vomiting) or the positive rapid antibody covid-19 test, or the chest x-ray findings, or the abnormality of complete blood count (CBC) especially NLR >5.8 (Figure 1). The inclusion criteria of this study is all pregnant women suspected of COVID-19 (based on our screening methods) that delivered in Universitas Airlangga hospital during study periods. There is no spesific exclusion criteria in this study. All participant were recruited for this study following informed consent. The final diagnosis of all subjects was then confirmed with the RT-PCR. The sample was taken from the nasopharyngeal and oropharyngeal swabs. The subject was divided into two groups based on the RT-PCR result as non-covid or covid groups.

**Figure 1.**
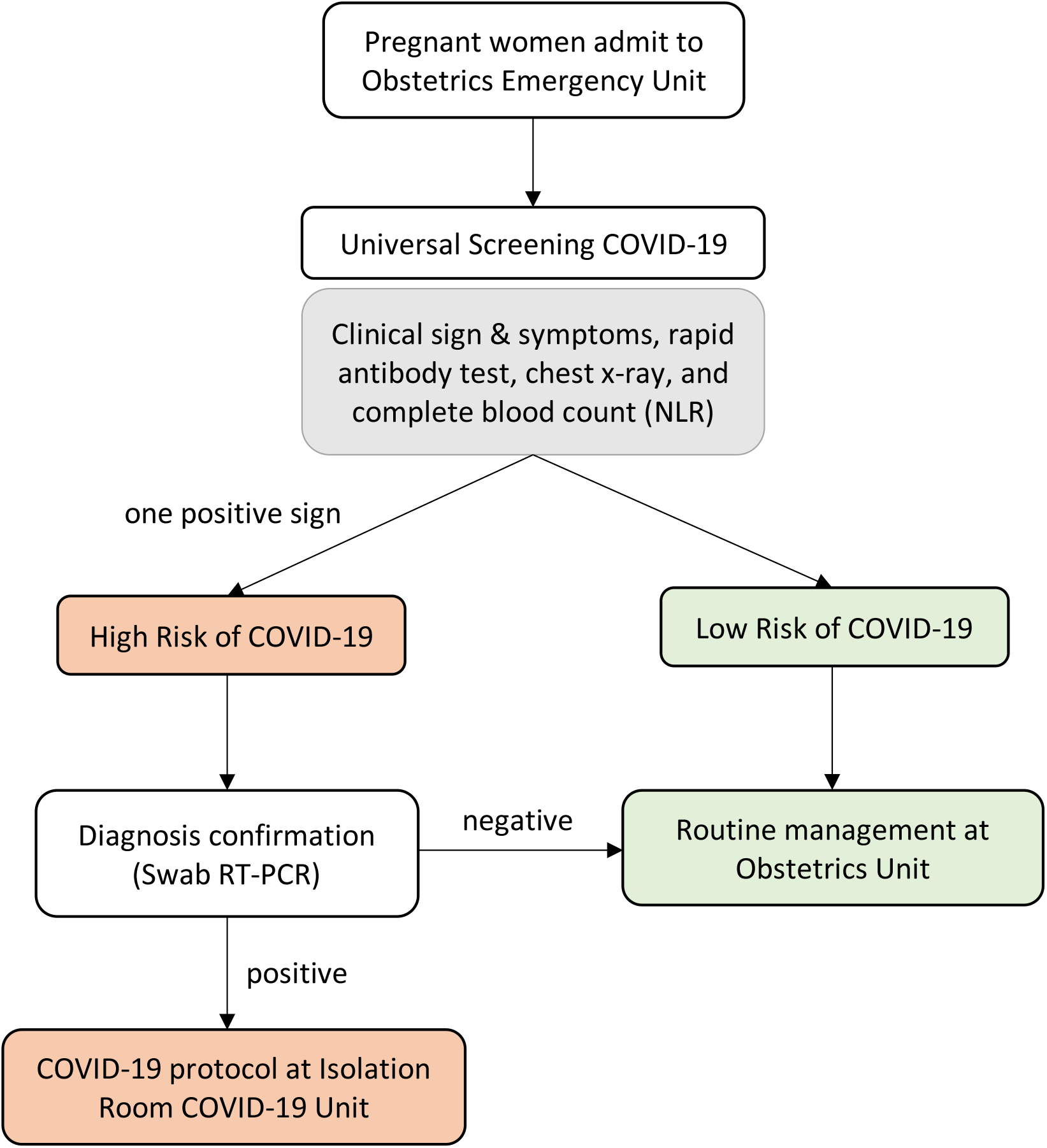
Algorithm of Screening of COVID-19 in Pregnant Women Admitted to Hospital. All pregnant women admit to the hospital will be screened for the COVID-19 infection using the screening methods consist of: sign & symptoms, rapid antibody test, chext x-ray and NLR > 5.8. The presence of one positive sign indicate a high risk COVID-19 women, and need an additional diagnosis methods (swab RT-PCR). Women with positive molecular test were managed using a COVID-19 protocol at isolation room, while the negatives were routinely managed in obstetrics unit.

The study’s primary outcomes were the diagnostic accuracy of the clinical sign, rapid antibody test, chest x-ray, NLR, and the combined clinical sign compared to the RT-PCR result. The combined clinical sign was defined as a presence of minimal one suspicious sign and symptoms of COVID-19. The presence of peripheral airspace shadowing and bilateral multilobular ground-glass opacities in chest x-ray was considered a suspicious sign of COVID-19. Rapid antibody test was performed using “One Step Rapid Test” - SARS-CoV-2 IgM/IgG Ab Rapid Test Kit (Qingdao Hightop Biotech Co., LTD). This test is used to detect the IgM and IgG antibodies to SARS-CoV 2 in human serum, plasma, or whole blood. The sensitivity and specificity of this test claimed by the company: IgM (83% & 97%) and IgG (93% & 97.5%) (17). Peripheral blood NLR (Neutrophil/Lymphocyte Ratio) has been shown by many studies as a good marker of systemic inflammation, likewise in COVID-19 infection (18). NLR was count by dividing Neutrophil count with Lymphocyte count from the Complete Blood Count examination. We used a cut-off value of NLR > 5.8 as a screening method of COVID-19.

### Ethical Consideration

This study has been approved by the Research Ethics Committee of Universitas Airlangga Hospital (Ref. No: 110/KEP/2021). All participant has understand the information of consent of this study including the objective, protocol, risk, and anonimity. All participant has signed the informed consent form before involved in the study.

### Statistical Analysis

The data was analyzed using IBM^@^ SPSS^©^ Statistics ver 25. Descriptive statistics was used first to determine the distribution normality of the numerical variables. The numerical variables with a normal distribution (hemoglobin, leucocytes, thrombocyte) were analyzed using an independent t-test value (table 1). The numerical variables with abnormal distribution (Neutrophil, Lymphocyte, NLR) was applied to the Mann Whitney test (table 1). A Chi-Square/ Fischer’s exact test was used to analyze the difference between categorical variables (table 1). Significant value is defined as p-value < 0.05. To evaluate the diagnostic accuracy of all the variables we measured: the sensitivity, specificity, positive likelihood ratio (PLR), negative likelihood ratio (NLR*), positive predictive value (PPV), negative predictive value (NPV), and overall accuracy (table 2).

**Table 1.** Screening and Diagnostic Test.

|  | Non Covid<br>n = 79 | Covid<br>n = 62 | P-value |
| --- | --- | --- | --- |
| Sign & Symptoms | 19 (24.1%) | 15 (24.2%) | 0.984 |
| Cough | 16 (20.3%) | 12 (19.4%) | 0.894 |
| Fever | 5 (6.3%) | 9 (14.5%) | 0.107 |
| Dyspneu | 6 (7.6%) | 8 (12.9%) | 0.295 |
| Sore throat | 4 (5.1%) | 3 (4.8%) | 0.951 |
| Myalgia | 0 | 1 (1.6%) | 0.440 |
| Nausea Vomiting | 2 (2.5%) | 4 (6.5%) | 0.405 |
| Rapid antibody test<br>status |  |  |  |
| Negative | 27 (35.1%) | 15 (27.3%) | 0.343 |
| Positive | 50 (64.9%) | 40 (72.7%) |  |
| Rapid antibody test<br>status |  |  |  |
| Negative | 27 (34.2%) | 15 (24.2%) | 0.245 |
| Ig M positive | 10 (12.7%) | 7 (11.3%) |  |
| IgG positive | 16 (20.3%) | 13 (21%) |  |
| Ig M & Ig G positive | 24 (30.4%) | 20 (32.3%) |  |
| Not examined | 2 (2.5%) | 7 (11.3%) |  |
| Chest x-ray |  |  |  |
| Normal | 47 (59.5%) | 35 (56.5%) | 0.142 |
| Abnormal | 32 (40.5%) | 24 (38.7%) |  |
| Not performed | 0 | 3 (4.8%) |  |
| NLR |  |  |  |
| NLR < 5.8 | 54 (72%) | 34 (58.6%) | 0.106 |
| NLR ≥ 5.8 | 21 (28%) | 24 (41.4%) |  |
| Hemoglobin | 11.34 ± 1.48 | 10.83 ± 1.80 | 0.072 |
| Leucocyte<sup>a</sup> | 11270 ± 3500 | 10552 ± 5002 | 0.327 |
| Trombocyte<sup>a</sup> | 279835 ± 83053 | 289306 ± 80894 | 0.504 |
| Neutrophil | 76.1 (9.8) | 76.6 (11.75) | 0.217 |
| Lymphocyte<sup>s</sup> | 15.5 (7.15) | 15.8 (9.33) | 0.309 |
| NLR<sup>B</sup> | 4.88 (2.55) | 4.94 (4.15) | 0.226 |

This is the screening and diagnostic test comparison between Covid and non covid groups, consist of sign and symptoms, rapid antibody test status, laboratory result, and chest x ray. The categorical variables were analyzed using a Chi-square and Fischers exact test, while the numerical variables were analyzed using an independent t-test.
Note: <sup>a</sup> indicates data with normal distribution, present with mean ± SD. <sup>b</sup> indicate data with abnormal distribution, present with median (interquartile range).

**Table 2.** Diagnostic accuracy of Clinical Sign & Symptoms, Laboratory, Rapid Antibody test, and Chest X-ray.

|  | Sensitivity<br>% (95%<br>CI) | Specificity<br>% (95%<br>CI) | Positive<br>Likelihood<br>Ratio | Negative<br>Likelihood<br>Ratio | Positive<br>Predictive<br>Value<br>% (95%<br>CI) | Negative<br>Predictive<br>Value<br>% (95%<br>CI) | Accuracy<br>% (95%<br>CI) |
| --- | --- | --- | --- | --- | --- | --- | --- |
| Cough | 19.35<br>(10.42-31.37) | 79.75<br>(69.20-87.96) | 0.96<br>(0.49-1.87) | 1.01<br>(0.86-1.19) | 3.73<br>(1.94-7.05) | 96.06<br>(95.38-96.64) | 77.39<br>(69.59-84) |
| Fever | 14.52<br>(6.86-25.78) | 93.67<br>(85.84-97.91) | 2.29<br>(0.81-6.50) | 0.91<br>(0.81-1.03) | 8.52<br>(3.18-20.87) | 96.43<br>(96-96.81) | 90.58<br>(84.51-94.85) |
| Dyspnea | 12.9<br>(5.74-23.85) | 92.41<br>(84.2-97.16) | 1.7<br>(0.62-4.64) | 0.94<br>(0.84-1.06) | 6.45<br>(2.46-15.85) | 96.32<br>(95.85-96.70) | 89.3<br>(82.99-93.88) |
| Sore<br>throat | 4.84<br>(1.01-13.5) | 94.94<br>(87.54-98.60) | 0.96<br>(0.22-4.11) | 1.00<br>(0.93-1.08) | 3.73<br>(0.89-14.30) | 96.09<br>(95.8-96.37) | 91.42<br>(85.52-95.48) |
| Nausea<br>vomiting | 6.45<br>(1.79-15.7) | 97.47<br>(91.15-99.69) | 2.55<br>(0.48-13.46) | 0.96<br>(0.89-1.03) | 9.37<br>(1.92-35.33) | 96.25<br>(95.97-96.51) | 93.92<br>(88.61-97.25) |
| Combined Sign &<br>symptoms | 24.19<br>(14.22-36.74) | 75.95<br>(65.02-84.86) | 1.01<br>(0.56-1.81) | 1.00<br>(0.83-1.20) | 3.92<br>(2.21-6.86) | 96.11<br>(95.34-96.75) | 65.87<br>(65.87-80.96) |
| Rapid<br>antibody<br>test | 72.73<br>(59.04-83.86) | 35.06<br>(24.53-46.78) | 1.12<br>(0.89-1.41) | 0.78<br>(0.46-1.32) | 4.35<br>(3.48-5.41) | 96.94<br>(94.92-98.17) | 36.53<br>(28.33-45.36) |
| Chest X-Ray | 40.68<br>(28.07-54.25) | 59.45<br>(47.85-70.40) | 1.00<br>(0.67-1.51) | 1.00<br>(0.75-1.32) | 3.92<br>(2.64-5.77) | 96.11<br>(94.92-97.03) | 58.76<br>(50.07-67.07) |
| NLR ≥<br>5.8 | 41.38<br>(28.60-55.07) | 72.00<br>(60.44-81.76) | 1.48<br>(0.92-2.38) | 0.81<br>(0.63-1.05) | 5.66<br>(3.60-8.80) | 96.80<br>(95.90-97.51) | 70.81<br>(62.30-78.36) |

This is the diagnostic accuracy parameter for each individual variables to detect covid-19, consists of: sign & symptoms, rapid antibody test, chest x-ray, and NLR. All parameter were performed using diagnostic accuracy test to evaluate the sensitivity, specificity, positive likelihood ratio, negative likelihood ratio, positive predictive value, negative predictive value and diagnostic accuracy.

## Result

A total of 141 suspected covid-19 cases in pregnancy were recruited during this study period. Sixty-two subjects were confirmed as COVID-19 cases (43.9%) and seventy-nine subjects as non-COVID-19 cases (56.1%). The summary of screening and diagnostic results between both groups can be seen in table 1. The presence of combined signs and symptoms did not differ between both groups. We also compared and analyzed each sign & symptoms individually between both groups (cough, fever, dyspnea, sore throat, myalgia, and nausea vomiting) and found no significant differences. The rapid antibody test once again fails to show any difference between both groups. 2,5% and 11.3% of patients from non-covid-19 and covid-19 groups did not have a rapid antibody test examined. Furthermore, the abnormalities of chest x-ray and NLR >5.8 is not different between both groups. Additionally, the complete blood count (hemoglobin, leucocyte, thrombocyte, neutrophil, lymphocyte, and NLR) mean/median value is not significantly different between these two groups.

The diagnostic accuracy of the clinical sign-symptoms, laboratory test, rapid antibody covid-19 test, and a chest x-ray was further analyzed (Table 2). Rapid antibody test has the highest sensitivity compared to other modalities (72.73%), while on the contrary, sore throat has the lowest one (4.84%). The clinical manifestation of nausea, vomiting, sore throat, fever, and dyspnea have the highest specificity compared to other examinations (97.47%, 94.94%, 93.67%, and 92.41%). The highest PLR was found in the existence of fever (2.29). On the other hand, the highest NLR was found in cough, sore throat, combined sign-symptoms, and chest x-ray (1.00-1.01). The presence of nausea, vomiting, and fever has the highest positive predictive value (9.37% and 8.52%), although the actual value is deficient. This finding was in contrast with the negative predictive value measurement, which showed a relatively high percentage in all screening-diagnostic modalities (96.0 - 96.9%). The overall diagnostic accuracy of these modalities was varied (36.53-93.92), with the highest one include nausea, vomiting (93.92%), sore throat (91.42%), and fever (90.58%). We analyze the NLR value’s ROC curve to predict COVID-19 infection in pregnant women (figure 2), and the Area Under Curve (AUC) value is 0.561. With the cut-off value that we used (5.8), the sensitivity and specificity from the ROC curve were only 37.9% and 30.7%.

**Figure 2.**
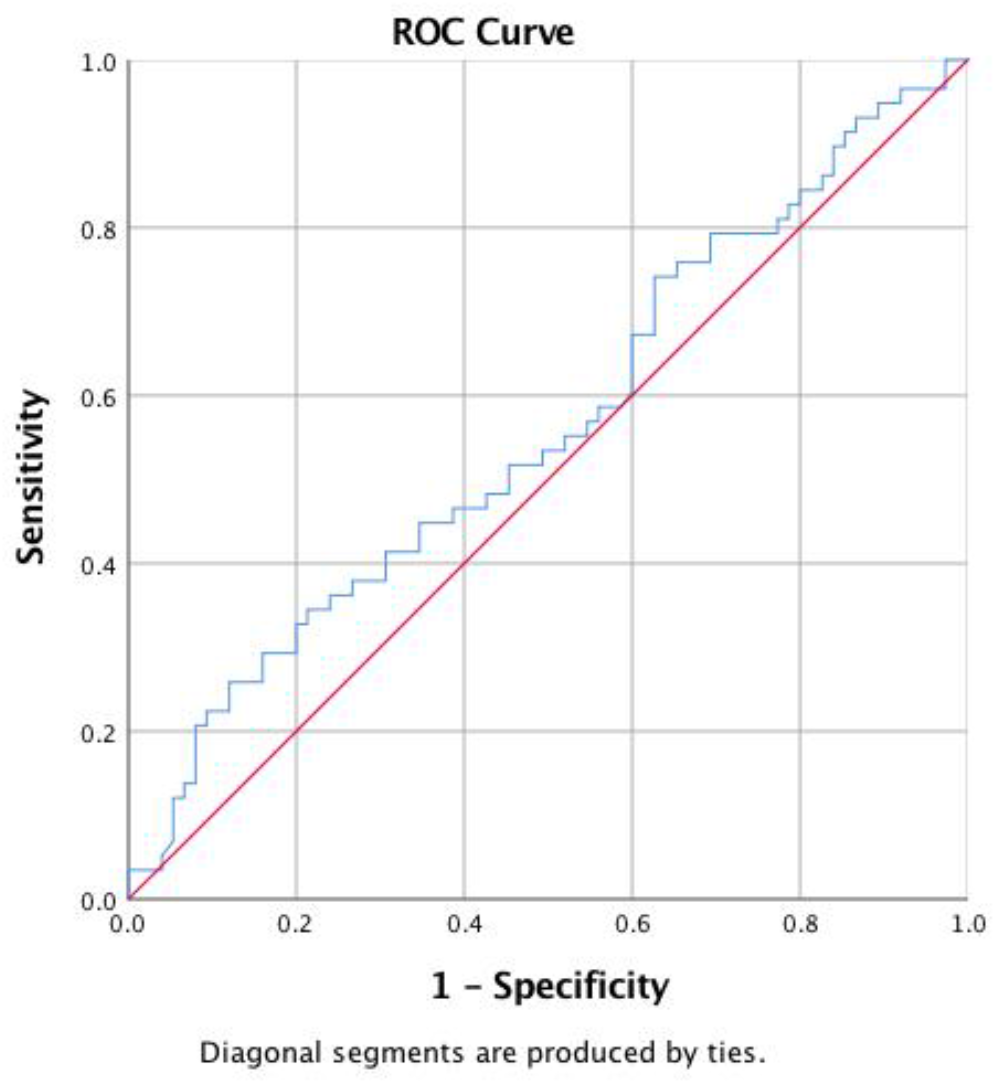
ROC Curve of The NLR to predict COVID-19 infection in pregnant women. NLR > 5.8 has an AUC of 5.61 to predict the COVID-19 infection in pregnant women. The sensitivity and specificity from the ROC curve were only 37.9% and 30.7%.

## Discussion

This study point that the screening methods of COVID-19 have various diagnostic accuracy (DOR: 36.53-93.92). The presence of individual signs and symptoms has the highest diagnostic accuracy (with the order: nausea, vomiting, sore throat, fever, dyspnea, and cough) followed by NLR, chest x-ray, and serological test. However, the diagnostic accuracy of combined sign and symptoms occupy the second rank after NLR. In pregnancy, this becomes a problem since most pregnant women with COVID-19 have no clinical symptoms (3,19,20). A meta-analysis study including 96.604 participants compared pregnant women with and without COVID-19 infection from the universal screening, show that three-quarter of 162 infected pregnant women was asymptomatic (74% from 11 studies) (21). A similar result is also found in this study: 75% of pregnant women with COVID-19 has no symptoms when admitted to the hospital. The most significant and symptoms found in this study are cough, fever, and dyspnea, in line with many studies (21–26). Unfortunately, in this study, we did not evaluate anosmia and ageusia symptoms, which were later known as an essential clinical sign of SARS-CoV-2 infection (27). The combined clinical symptoms, in general not appear to be an adequate screening method, considering the low sensitivity (24.19%) and PPV (3.92%). On the other hand, clinical symptoms have a reasonable specificity and NPV: the specificity for individual symptoms: >75%, for combined symptoms: >90%, and NPV >96%. In other words, the combined clinical sign and symptoms have a low false-positive rate for diagnosing the SARS-CoV-2 infection. So that in pregnant women with high suspicion of COVID-19 infection, if we found one of the clinical symptoms, it is most likely infected with SARS-CoV-2. In general, the presence of one symptom has a moderate diagnostic accuracy (DOR: 65.87). This study indicates that clinicians can utilize the sign and symptoms to rule in pregnant women with suspicion of COVID-19.

The finding of infiltrate on chest x-ray in this study shows a moderate diagnostic accuracy of COVID-19 in pregnant women. CT chest is usually preferred in diagnostic of COVID-19 compared to a chest x-ray. Many guidelines recommend using CT chest as a primary diagnostic tool in pregnant women with suspicion of COVID-19 in a high endemic area (28). Chest CT has known to have a high sensitivity to diagnose COVID-19 (29). Unfortunately, in low-middle income countries, the chest CT is often unavailable; therefore, a chest x-ray can be utilized as an alternative substitute. This study shows a moderate result of sensitivity, specificity, and NPV of a chest x-ray to diagnose COVID-19 infection (40.68%, 59.45% dan 96.11%). This method can be used as an early screening method in pregnant women with suspicion of COVID-19. A study from India reports the use of a*rtificial intelligence* assisted chest x-ray examination to increase the detection accuracy of COVID-19 (90.5% with sensitivity 100%) (30). These findings will be beneficial in countries with limited resources to increase the screening and detection of COVID-19 by using chest x-ray in high-risk pregnant women. Therefore our national Obgyn society (POGI) has released an official recommendation to use chest x-ray as an alternative option for screening COVID-19 in pregnant women if the CT chest is unavailable (9).

The laboratory result between both groups does not significantly differ in hemoglobin, leukocyte, thrombocyte, neutrophil, lymphocyte, and NLR value. Many studies and meta-analyses indicate that COVID-19 infection in an adult manifests as decreased leukocyte (otherwise, in severe cases, leucocytosis), thrombocytopenia, decrease hemoglobin, neutrophilia, lymphopenia, and increase NLR value (21,23,31–34). Unfortunately, these changes are often found inconsistent in pregnant women. Pregnancy causes essential physiological changes, which lead to insignificant laboratory values changes in SARS-CoV-2 infection. For example, in the first-second trimester, leukocyte and lymphocyte values increase before finally decreasing in the third trimester (35). Neutrophilia can be found in the second-third trimester caused by disruption of neutrophil apoptosis during pregnancy (36). The platelet value will also decrease in the third trimester, known as gestational thrombocytopenia (35). Study about laboratory changes in pregnancy is still minimal and provide inconsistent result. Liu et al. study shows a leukocytosis in half of the infected pregnant women; on the other hand, Chen et al. show that most pregnant women with COVID-19 show a decrease leukocyte value (10,14). Neutrophil value is found to increase in pregnant women with COVID-19 in Liu et al. study, but Li et al. study shows an opposite result (14,37).

NLR has been known as a consistent marker to predict the progression of COVID-19 disease. The increased level of NLR appears in a critically ill or severe COVID-19 case (18). A meta-analysis study, which includes 1579 participants, shows that NLR has excellent overall sensitivity, specificity, and AUC in predicting the severity of COVID-19 (0.78, 0.78, dan 0.85) (18). In this study, we use a high cut off value of NLR (> 5.8) as a screening method of COVID-19 in pregnancy. The result shows a lower AUC value compared to Li et al. study (0.561 vs 0.85) (18). Our study shows that NLR > 5.8 has the highest diagnostic accuracy of other screening methods (DOR: 70.81). The sensitivity of NLR in our study is lower than the meta-analysis by Li et al. (41.38% vs 78%), but the specificity is similar (72 vs 78%) (18). COVID-19 in pregnancy causes an excessive inflammatory reaction, leading to cytokine storms and poor clinical outcomes. The presence of exaggerated inflammatory reaction and immune system suppression causes a progressively decreased lymphocyte number and increased neutrophil. A neutrophil is a proinflammatory cell triggered by proinflammatory cytokine produced by SARS-CoV-2 such as IL-1, IL-6, and IL 8. On the other hand, this over inflammatory reaction will suppress lymphocyte production of CD4 and CD8 T cells (38,39). Increased NLR level is one of the peripheral blood examinations which indicate an inflammatory reaction. Therefore pregnancy with COVID-19 will show an increased level of NLR. However, this is not found in our study; the mean NLR value between infected and non-infected pregnant women is not statistically different. The possible explanation for this finding may be related to the majority of our COVID-19 patient was mildly affected. Some studies have shown that the NLR value in severe COVID-19 patients was higher than the mild cases (18,40,41).

Rapid antibody serological test from our study has a relatively low detection accuracy. The sensitivity of the rapid antibody test is moderate, but it has a low specificity. This finding is not following a study result in Italia (42). In a meta-analysis study which include 29.842 test, rapid antibody test with ELISA method show an overall good sensitivity and specificity (sens: 84.3%, 95% CI: 75.6-90.9% & specs: 99.7%, 95% CI: 99-100%) (16). The rapid antibody test is not sufficient to be used as an early screening method of COVID-19 in pregnancy (42). With a specificity of 35%, there is a possibility of 65% of pregnant women infected by SARS-CoV-2 get a negative test result (false negative), so that this test will lead to many misdiagnoses. One of the possible explanations of the serological test’s low accuracy in our study may be related to the timing of blood sampling. The majority of the test was performed in the acute early stage of the infection when the patient was admitted to the hospital. The sensitivity and specificity of antibody tests will be much higher in the third week of infection (16,43). The rapid antibody test is more appropriate as an epidemiological surveillance tool of the disease in the general population and to diagnose the patients’ immunity status (infection phase) (16,42–44). An antibody test can not be used as a single screening or diagnostic method for COVID-19 infection and need a combination with clinical sign, laboratory, imaging, and RT=PCR test. The serological test can still be used in an algorithm of COVID-19 management to evaluate the recovery phase of mildly affected patients whose in self-isolation (surveillance). The negative IgM SARS-CoV-2 from the serological test may be used as an indicator of discontinuing the self-isolation periods (43). Recently serological antigen test of SARS-CoV-2 has been developed, which has a higher sensitivity, specificity, and accuracy than the serological antibody test in an early phase of infection (45).

This study’s limitation is that the study population was all pregnant women with suspicion of COVID-19 infection. This matter will cause an unclear changes in clinical signs, laboratory, and additional examination because the control group was also a high-risk COVID-19 population. Furthermore, we cannot evaluate this screening method’s accuracy in the population of normal pregnant women (high and low risk) because of this inclusion criteria. We assume that the accuracy of this screening methods will be higher in the general population study.

In a low middle income countries where the RT-PCR test was limited, the goverment can used the combined screening methods consist of clinical sign and symptoms, serological test, chest x-ray, and NLR to classify pregnant women with high risk of COVID-19 infection. Pregnant women with at least one sign from screening method can be further diagnose with RT-PCR test. This approach can be used in a limited resources hospital or health services in low middle income countries.

## Conclusion

This study found a moderate accuracy of a single parameter of the screening methods to detect COVID-19 in pregnant women. A combination of multiple parameters (such as clinical signs & symptoms, serological test, chest x-ray, and NLR) could help classify pregnant women with a high risk of COVID-19 in the limited resources area. Pregnant women with positive signs from the screening methods can be further diagnosed with the RT-PCR test’s gold standard. This practice will help to reduce the massive burden of RT-PCR examination in an area/country without sufficient resources.

## Data Availability

The data that support the findings of this study are available from Universitas Airlangga Hospital. Restrictions apply to the availability of these data, which were used under license for this study. Data are available from the authors with the permission of Universitas Airlangga Hospital.

## Acknowledgements

We thank Prof. Nasronudin, MD, Ph.D., as the Director of Universitas Airlangga Hospital and all directors to support this study. We thank Brahmana Askandar Tjokroprawiro, MD, Ph.D. as the Head of Department Obstetrics & Gynaecology Faculty of Medicine Universitas Airlangga and all the staff for their valuable input, comments, opinion, advice, and support in this study. We thank all the research team for their enormous effort in collecting data for this study. We thank all the patients for their participation in this study.

